# Modeling analysis of COVID 19-related delays in colorectal cancer screening on simulated clinical outcomes

**DOI:** 10.1101/2022.11.17.22282447

**Authors:** Lesley-Ann Miller Wilson, Vahab Vahdat, Durado Brooks, Paul Limburg

**Affiliations:** Exact Sciences, 5505 Endeavor Lane, Madison WI, 53719

**Keywords:** COVID-19, early detection of cancer, pandemics, risk factors, prevention & control

## Abstract

**Objective:** Colorectal cancer (CRC) screening disruptions observed during the COVID-19 pandemic put patients at risk for more advanced-stage disease when diagnosed. This budget impact simulation model assessed increased use of multi-target stool DNA [mt-sDNA] or fecal immunochemical [FIT] tests to offset disruption in colonoscopy screening due to COVID-19 in adults at average-risk for CRC, from a United States payer perspective

**Main outcomes and measures:** Compared to the base case (S0; 85% colonoscopy and 15% non-invasive tests), the estimated number of missed CRCs and advanced adenomas (AAs) were determined for four COVID-19-affected screening scenarios: S1, 9 months of CRC screening at 50% capacity, followed by 21 months at 75% capacity; S2, S1 followed by increasing stool-based testing by an average of 10% over 3-years; S3, 18 months of CRC screening at 50% capacity, followed by 12 months of 75% capacity; and S4, S3 followed by increasing stool-based testing by an average of 13% over 3-years.

**Results:** Increasing the proportional use of mt-sDNA improved AA detection by 6.0% (Scenario 2 versus 1) to 8.4% (Scenario 4 versus 3) and decreased the number of missed CRCs by 15.1% to 17.3%, respectively. Increasing FIT utilization improved the detection of AAs by 3.3% (Scenario 2 versus 1) to 4.6% (Scenario 4 versus 3) and decreased the number of missed CRCs by 12.9% to 14.9%, respectively. Across all scenarios, the number of AAs detected was higher for mt-sDNA than for FIT, and the number of missed CRCs was lower for mt-sDNA than for FIT.

**Conclusions and relevance:** Using home-based stool tests for average-risk CRC screening can mitigate the consequences of reduced colonoscopy screening resulting from the COVID-19 pandemic. Use of mt-sDNA led to fewer missed CRCs and more AAs detected, compared to FIT.

**Key Points:** 

**Question:** What is the impact of increasing the use of stool-based screening tests for colorectal cancer (CRC) during the COVID-19 pandemic in the United States?

**Findings:** In this simulation model, increasing the use of stool-based screening tests increased the number of advanced adenomas detected and decreased the number of missed CRC cases. Use of multi-target stool DNA (mt-sDNA) resulted in a higher number of advanced adenomas detected and a lower number of missed CRC cases compared to fecal immunochemical testing (FIT).

**Meaning:** Increased use of mt-sDNA led to fewer missed CRC cases and more advanced adenomas detected, compared to FIT, when simulating reduced colonoscopy screening resulting from the COVID-19 pandemic.

## INTRODUCTION

Colorectal cancer (CRC) is the second most common cause of cancer deaths in the United States (US).1 Though CRC is most frequently diagnosed among persons aged 65 to 74 years, nearly 7% of new CRC cases occur in persons younger than 50 years1 and the incidence of cancer in those aged 40 – 49 years has increased by 15% from 2000 – 2002 to 2014 – 2016.2 Due to the existence of a screen-detectable precursor lesion, the long duration of disease manifestation and the high mortality rate associated with advanced-stage disease, CRC screening represents an attractive opportunity for improving public health outcomes. Further, screening for CRC is relatively simple and reasonable treatment options exist for those diagnosed.^3^ To that end, the United States Preventive Services Task Force (USPSTF) has recently updated their guidelines to recommend that, in addition to those over the age of 50 years, adults aged 45 to 49 years should be screened.^4^

CRC screening is associated with decreases in CRC incidence and mortality.^5^ However, in order to fully realize the benefits of CRC screening, individuals with normal stool-based screening tests need to repeat screening at pre-specified regular intervals and those with abnormal test results need additional evaluation via follow-up (and potentially ongoing surveillance) colonoscopy.^6^ The coronavirus disease 2019 (COVID-19) pandemic disrupted cancer screening and treatment activities, with cancer screening programs paused for varying lengths of time.^7^ This disruption was most evident for CRC screening options that require individuals to be present at health care facilities (e.g., colonoscopy), and has been projected to result in substantial increases in avoidable cancer deaths due to diagnostic delays.8 Modeling studies have estimated that the screening and treatment delay due to COVID-19 pandemic will lead to an increase of 1% of cancer-specific deaths over a period of 10 years, many of which would have been otherwise preventable.9

A previous analysis explored how expanding the use of home-based options for CRC screening could favorably affect participation during the COVID-19 pandemic.10 This study, however, only considered increasing participation with fecal immunochemical test (FIT)-based screening; did not include other accessible non-invasive screening tests, such as multi-target stool DNA (mt-sDNA); and, did not consider a start age of screening at 45 years old, as per the latest USPSTF guidelines.^4^ The currently reported analyses expand on previous publications and estimate the degree to which increasing non-invasive CRC stool-based screening with mt-sDNA or FIT for CRC, during a public health crisis such as the COVID-19 pandemic, impacts clinical outcomes.

## METHODS

### Overview

A previously developed Markov simulation model^11^ was adapted to estimate the degree to which COVID-19 impacted colorectal cancer screening participation and the associated contribution of this decline in CRC screening on outcomes from a payer perspective. While details of the original model are reported elsewhere^11^, briefly, the model was adapted to evaluate the screening cohort in annual cycles to assess the clinical consequences of CRC screening with different modalities at varying utilizations levels. **Table 1** shows the screening performance characteristics by modality. Annual risk reductions for both mt-sDNA and FIT were used from a recently published study by the USPSTF.^12^ Adverse event rates associated with colonoscopy were the same as those in the previously published simulation model.^11^ This modeling study follows the methods of budget impact analysis reporting, as outlined by the International Society for Pharmacoeconomics and Outcomes Research guidelines for good practice; this paper, however, only reports on the clinical outcomes findings.^13,14^

**Table 1.** Model input values and data sources.

|  | Sensitivity |  | Specificity |
| --- | --- | --- | --- |
| Age band | CRC | Advanced adenomas (≥10mm) |  |
| mt-sDNA<sup>15</sup> |  |  |  |
| 40-44 | 92.3% | 38.0% | 92.2% |
| 45-49 | 92.3% | 38.0% | 92.2% |
| 50-54 | 92.3% | 38.0% | 92.2% |
| 55-59 | 92.3% | 38.0% | 92.2% |
| 60-64 | 92.3% | 42.1% | 89.0% |
| 65-69 | 92.3% | 41.5% | 85.7% |
| 70-74 | 92.3% | 46.8% | 82.5% |
| 75-79 | 92.3% | 46.8% | 77.8% |
| 80-85 | 92.3% | 46.7% | 77.9% |
| FIT<sup>16</sup> |  |  |  |
| All | 73.8% | 23.8% | 94.9% |
| Colonoscopy/Follow-up Colonoscopy<sup>12</sup> |  |  |  |
| All | 95.0% | 95.0% | 86.0% |

CRC: colorectal cancer; FIT: fecal immunochemical test; mt-sDNA: multi-target stool DNA

### Population and setting

The budget impact model estimated CRC outcomes over a 3-year time horizon, using the population estimates as outlined in the base case analysis **(Figure 1)**. Using an estimated population of 326,687,501 people in the US, and based on calculations from US Census data, 29% of people are aged 50 – 75 years and are considered eligible for screening.^17^ Of these, 67% are considered up to date with screening, 33% are eligible for screening, and 5.1% are estimated to complete screening in each of the three years.^18^ The model assumes that all individuals that completed screening were at average risk.^19^

**Figure 1.**
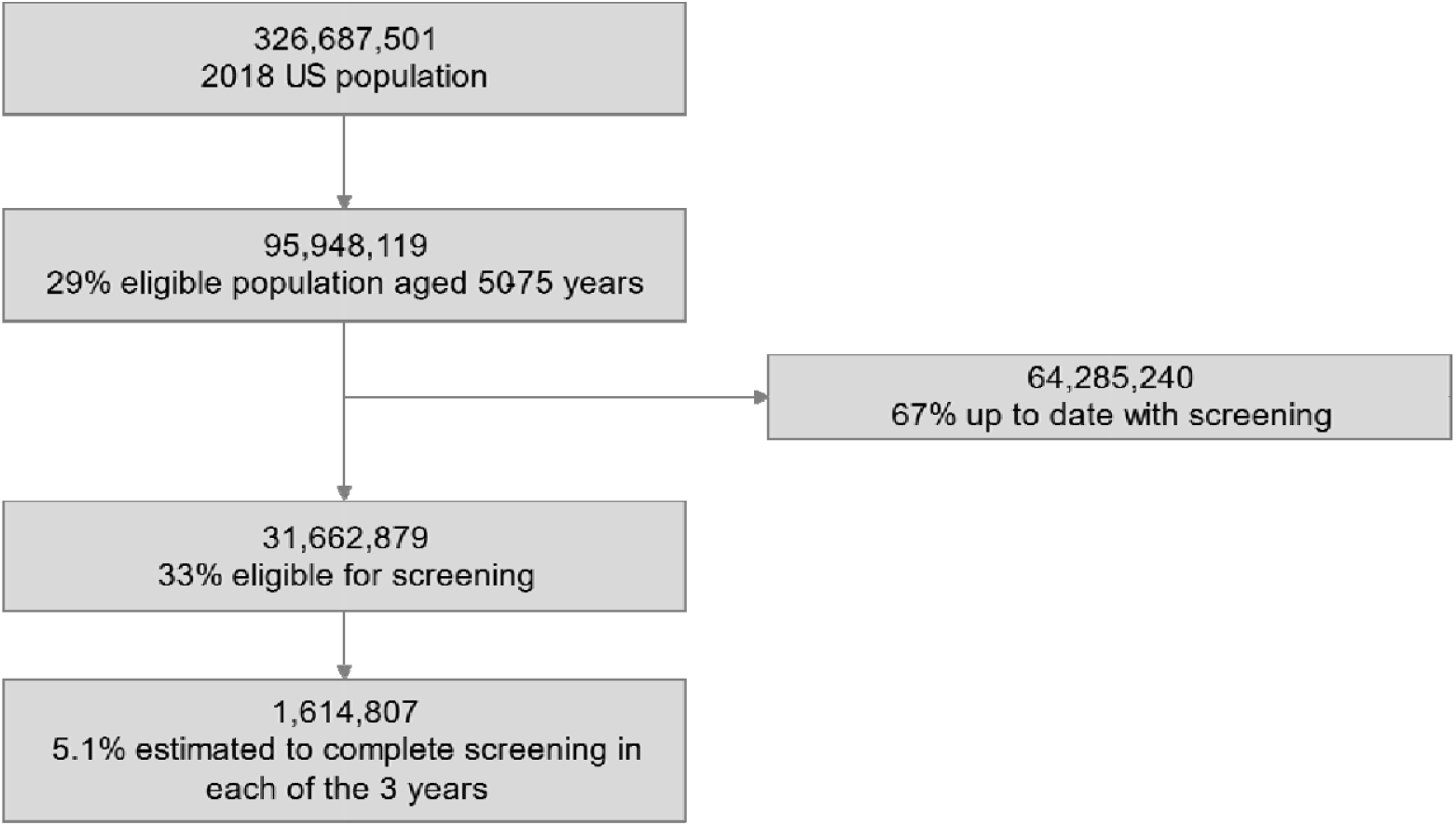
Model patient population flow, base case analysis (adapted from Issaka et al.^10^) US: United States

Two screening strategies were considered: a) colonoscopy and mt-sDNA; and b) colonoscopy and FIT. Age-specific risk profiles were assigned for incidence and prevalence of CRC and adenomas, by dividing the modeled population into 5-year age bands. Relative risk reductions were determined by screening prevalence and age band. CRC stages by screening modality were taken from the literature (**Table S1)**.

### Screening scenarios

In the base case scenario (S0), screening is assumed to continue under normal circumstances, that is, 85% of screening tests occur with colonoscopy and 15% occur using a non-invasive screening strategy. This status quo scenario is based on historical ACS data.^18^ Four COVID-19 reduced screening strategy scenarios were then explored over the 3-year time horizon (**Figure 2)**. In Scenario 1, compared to the base case scenario, there was an assumed reduction in CRC screening of 50% for nine months, followed by 21 months of CRC screenings at 75% of the base case levels. Scenario 2 is equivalent to Scenario 1, however, an increase in CRC screening of an average 10.0% via the use of non-invasive stool tests (mt-sDNA or FIT) over 3-years is explored to compensate for reduced colonoscopy screening (**Table S2**). Scenario 3 explored a longer return to screening (reflecting a prolonged impact of COVID-19), with a reduction in colonoscopy screening of 50% over 18 months, followed by 12 months of CRC screening at 75% return to normal. Scenario 4 explored Scenario 3 with an average 13.0% increase in the use of non-invasive stool tests (mt-sDNA or FIT) to compensate for reduced colonoscopy screening. The yearly proportion of the population being screened, by modality, is presented in **Table S2**.

**Figure 2.**
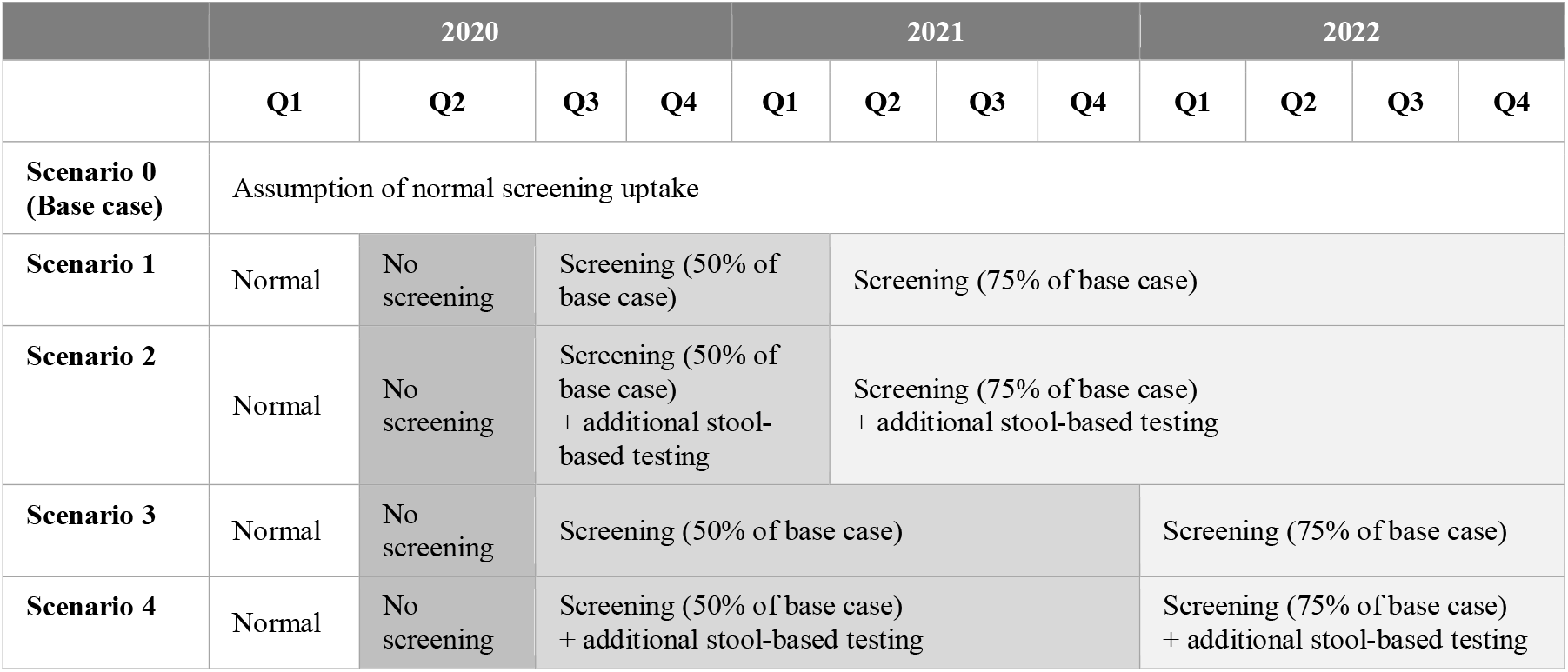
Modeled CRC screening scenarios.

### Outcomes

The number of patients screened, the number of advanced adenomas (AAs) detected and the number of missed CRC cases.

### Sensitivity analyses

In accordance with the most recent USPSTF guidelines, which recommend initiation of average-risk CRC screening at age 45 years, the impact of COVID-19-related delays was modeled over the age range of 45-75 years in sensitivity analyses (**Figure S1**).

## RESULTS

### mt-sDNA

Compared to all other scenarios, the base case scenario (S0), where there are no disruptions to screening due to the COVID-19 pandemic, had the lowest number of missed CRC cases and the highest number of AAs detected (**Table 2**). The highest number of AAs detected (and consequently the lowest number of missed CRC cases) was observed when indicated screening was not delayed, and screening was supplemented with increased mt-sDNA testing (Scenario 2). Compared to Scenario 1, Scenario 2 (increasing the number of mt-sDNA tests) increased the number of AAs detected and decreased the number of missed CRC cases. Among the four COVID-19 screening scenarios, Scenario 3 (prolonged reduced screening due to COVID-19) had the highest number of missed CRC cases and the lowest number of AAs detected; however, increased screening with mt-sDNA resulted in a lower number of missed CRC cases (Scenario 4).

**Table 2.** Total 3-year outcomes for COVID-19 screening scenarios: mt-sDNA.

| Clinical Outcomes | Scenario 0<br>(Base case) | Scenario 1 | Scenario 2 | Scenario 3 | Scenario 4 |
| --- | --- | --- | --- | --- | --- |
| Total eligible population during screening window | 4,844,421 | 4,844,421 | 4,844,421 | 4,844,421 | 4,844,421 |
| Number of screening colonoscopies | 4,117,757 | 2,648,283 | 2,648,283 | 2,389,914 | 2,389,914 |
| Number with no screening due to COVID-19 | 0 | 1,711,695 | 1,227,253 | 2,018,509 | 1,404,882 |
| Number of follow-up colonoscopies | 63,114 | 57,638 | 115,276 | 51,874 | 124,882 |
| Number of advanced adenomas detected | 97,172 | 63,620 | 67,425 | 57,404 | 62,223 |
| **Number of missed CRC Cases** | **1,398** | **3,180** | **2,701** | **3,499** | **2,893** |

CRC: colorectal cancer; mt-sDNA: multi-target stool DN

### FIT

Compared to all other scenarios, when using FIT as the non-invasive screening option, the base case scenario (S0) had the lowest number of missed CRC cases and the highest number of AAs detected (**Table 3**). Compared to Scenario 1, when COVID-19 was not prolonged, and screening was supplemented with increased FIT testing (Scenario 2), the number of AAs detected was higher and the number of missed CRC cases was lower. Among the four COVID-19 screening scenarios, Scenario 3 (no increased screening with FIT and prolonged reduced screening due to COVID-19) had the highest number of missed CRC cases and lowest number of AAs detected. As observed with mt-sDNA, when increasing screening with FIT, the number of AAs detected increased and the number of missed CRC cases decreased (Scenario 4).

**Table 3.** Total 3-year outcomes for COVID-19 screening scenarios: FIT.

| Clinical Outcomes | Scenario 0<br>(Base case) | Scenario 1 | Scenario 2 | Scenario 3 | Scenario 4 |
| --- | --- | --- | --- | --- | --- |
| Total eligible population during screening window | 4,844,421 | 4,844,421 | 4,844,421 | 4,844,421 | 4,844,421 |
| Number of screening colonoscopies | 4,117,757 | 2,648,283 | 2,648,283 | 2,389,914 | 2,389,914 |
| Number with no screening due to COVID-19 | 0 | 1,711,695 | 1,277,253 | 2,018,509 | 1,404,882 |
| Number of follow-up colonoscopies | 27,013 | 27,706 | 55,412 | 24,935 | 60,030 |
| Number of advanced adenomas detected | 94,983 | 61,843 | 63,870 | 55,805 | 58,373 |
| **Number of missed CRC Cases** | **1,486** | **3,239** | **2,820** | **3,552** | **3,021** |

CRC: colorectal cancer; FIT: fecal immunochemical test

### mt-sDNA versus FIT

While both scenarios S2 and S4, compared to scenarios S1 and S3, respectively, resulted in increased number of AAs detected and fewer missed CRC cases, the magnitude of these outcomes differed. Increasing the use of mt-sDNA increased the number of AAs detected by 6.0-8.4% and decreased the number of missed CRC cases by 15.1-17.3%. Increasing FIT utilization increased the number of AAs detected by 3.3-4.6% and decreased the number of missed CRC cases by 12.9-14.9%. Across all scenarios, the number of CRC cases missed is lower for mt-sDNA than for FIT (**Figure 3A**) and the number of adenomas detected is higher for mt-sDNA than for FIT (**Figure 3B**).

**Figure 3.**
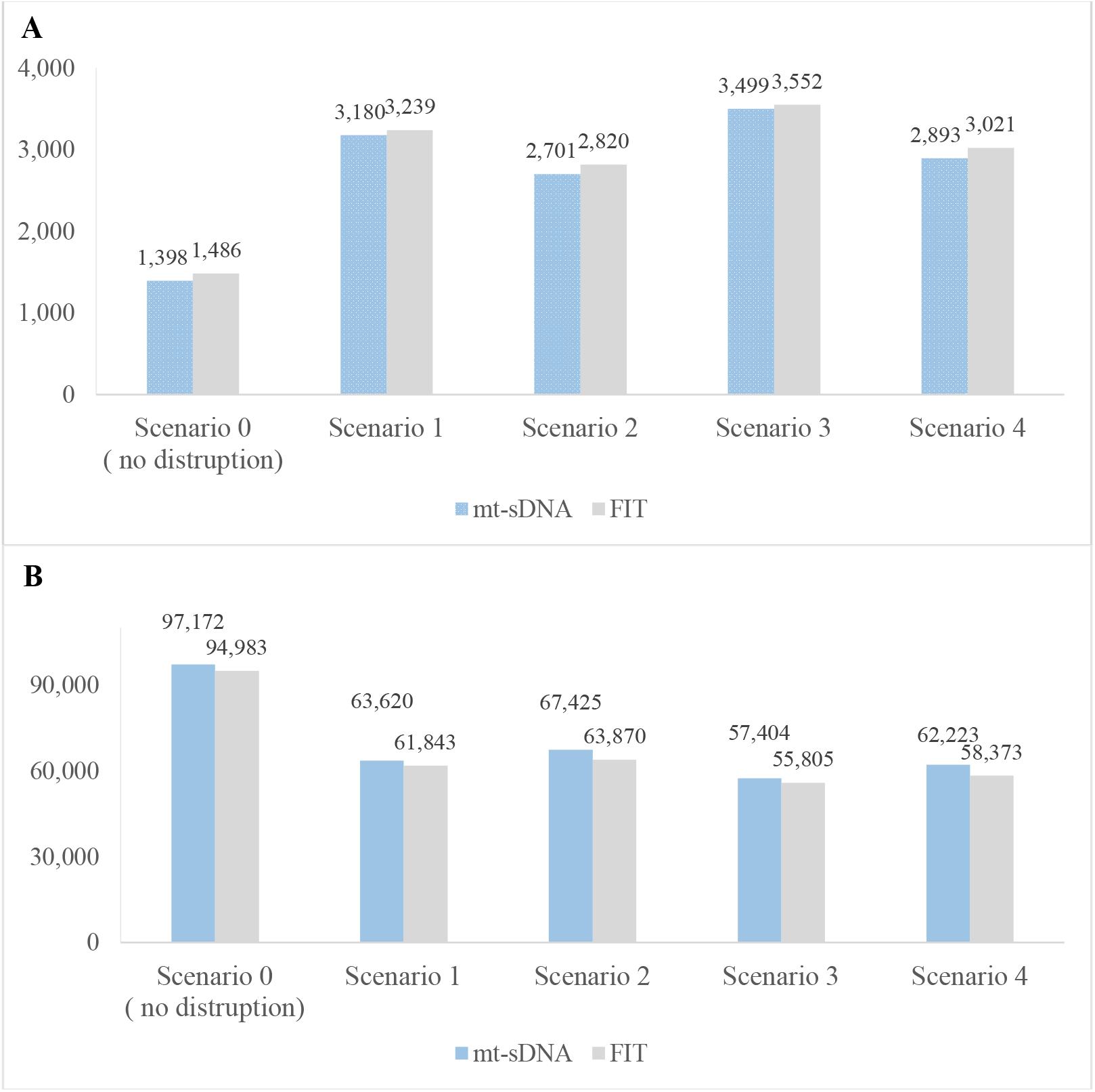
Clinical outcomes under COVID-19 scenarios, mt-sDNA versus FIT. (A) Number of missed CRC cases (B) Number of advanced adenomas detected. CRC: colorectal cancer; FIT: fecal immunochemical test; mt-sDNA: multi-target stool DNA

### Sensitivity analyses

As in the base case analysis, the number of missed CRC cases was lower and the number of AAs detected was higher with mt-sDNA than with FIT. Clinical outcomes when expanding the screening to those aged 45 to 75 years are presented in **Table S3** for mt-sDNA and **Table S4** for FIT. Given that 21% of those aged 45 – 49 reported being up to date with screening^18^, the remaining 79% are considered eligible for screening, resulting in an additional 16,338,278 individuals; the total eligible population in each of the three years for individuals 45 – 75 years is 2,450,609. While the direction of the results is similar to that of the base case analysis, there is an increase in the number of missed CRC cases for both mt-sDNA and FIT. The number of AAs detected, however, is increased for both mt-sDNA and FIT across all scenarios, as screening earlier allows the opportunity to detect more advanced adenomas.

## DISCUSSION

The COVID 19 pandemic has negatively impacted the delivery of healthcare across multiple areas, including CRC screening.^21,22^ While the full effects of the COVID-19 pandemic will not be known for years, the impact of diagnostic delays due to the interruption of screening have already disrupted timely cancer diagnosis.^23^ Further, the disruption in screening may potentially increase the mortality rate for otherwise generally preventable cancers^8^, with estimations of increases in cancer mortality of 2.5 cancer deaths per 100,000, without catching-up screening.^24^ This analysis demonstrated that interruptions in CRC screening due to the pandemic, compared to base case screening assumptions, increased the number of missed CRC cases and decreased the number of AAs detected. However, when increasing non-invasive screening tests to offset the reduction in colonoscopies, the number of missed CRC cases decreased, and the number of AAs detected increased. While this was observed for both mt-sDNA and FIT, mt-sDNA resulted in a greater reduction in number of missed CRC cases and a greater increase in AAs detected compared to FIT.

Other modeling studies have explored the impact of the COVID-19 pandemic on CRC screening clinical outcomes.^10^ Though Issaka et al. only explored one non-invasive stool-based test as an option to increase screening (FIT), they also found that under similar COVID-19 scenarios, increasing FIT use from 15% to 22% over a 3-year period resulted in an additional 2,715 CRC diagnoses.^10^ While it is difficult to compare across models due to different model structures and assumptions, our results found that increasing use of non-invasive screening tests, as seen in Scenario 2 and Scenario 4, decreased missed CRC cases by 15.1-17.3% and 12.9-14.9% for mt-sDNA and FIT, respectively. While our model assumed a prolonged return to normal^25^, other models have estimated that immediate catch-up on screening (assuming no delay in the return to normal) could minimise disruption and relative increase in colorectal cancer incidence and deaths to less than 0.1%.26

Our analysis also found a lower number of missed CRC cases and a higher number of AAs detected with mt-sDNA as compared to FIT. This is in due in part to the higher sensitivity of mt-sDNA versus FIT for cancer and advanced precancerous lesions.^27^ Further, while not explicitly modeled, mt-sDNA has test features that are compatible with telehealth consultation: it can be provider-ordered via an online portal; therefore no in-person visit is needed to order the test to be delivered a patient’s home; and, patient navigation is integrated with every test order, providing built-in support for those who need assistance. These characteristics of the mt-sDNA screening strategy may also contribute to improved outcomes relative to FIT screening.

Our model has some limitations that are worth noting. First, as with all simulation models, results are dependent on the generalizability and validity of the assumptions used within. The magnitude of the disruption due to COVID-19 on CRC screening over the long term remains difficult to project. Second, we assumed that while screening participation rates would be impacted by COVID-19, follow through would otherwise be unaffected. The completion of a follow-up colonoscopy after an abnormal stool-based screening test is required to fully complete the screening process. However, the model assumes all patients with an abnormal stool test result will complete the follow-up colonoscopy. Third, we modeled the impact of COVID-19 on CRC screening for a relatively short period (three years); in reality additional “waves” of COVID-19 outbreaks may continue to be experienced until vaccine effectiveness limits the spread.^28^ Fourth, this model assumes that there is capacity in the healthcare system to accommodate and interpret additional stool-based screening tests; the reality of this in a public health crisis is unknown.

## CONCLUSIONS

Increased utilization of home-based, non-invasive stool tests for average-risk CRC screening can help to mitigate the consequences of constrained capacity and acceptability of onsite screening options, such as colonoscopy, as the COVID-19 pandemic continues to evolve. In our modeling analyses, the use of mt-sDNA, with its integrated patient navigation, led to fewer missed CRC cases and more AAs detected, compared to FIT. Proactive strategies to increase the use of non-invasive stool-based screening, in particular mt-sDNA, may help increase CRC screening that declined as a result of the pandemic, contributing to the National Colorectal Cancer Roundtable goal of 80% screening rates across the US.^29^

## Data Availability

All data produced in the present study are simulated outcomes from the Exact Sciences' CRC BIM model.

## ACKNOWLEDGEMENTS

The authors would like to acknowledge Lianne Barnieh of the Maple Health Group for her medical writing services.

## AUTHOR CONTRIBUTIONS

All authors contributed to the conception and design of this study. All authors provided input on the development of the manuscript and approved the final version for submission.

## SUPPLEMENTARY MATERIALS

**Table S1.** Distribution of detected colorectal cancer by screening modality and stage.

| Screening modality | Stage I | Stage II | Stage III | Stage IV | Total |
| --- | --- | --- | --- | --- | --- |
| No screening<sup>12</sup> | 18% | 33% | 24% | 25% | 100% |
| mt-sDNA<sup>15</sup> | 44% | 36% | 15% | 5% | 100% |
| FIT<sup>20</sup> | 31% | 32% | 24% | 13% | 100% |
| Colonoscopy<sup>20</sup> | 43% | 23% | 27% | 8% | 100% |

**Table S2.** Yearly screening proportions (%)

|  | Scenario 0<br>(Base case) | Scenario 1 | Scenario 2 | Scenario 3 | Scenario 4 |
| --- | --- | --- | --- | --- | --- |
| **2020** |  |  |  |  |  |
| Colonoscopy | 85 | 42 | 42 | 42 | 42 |
| Stool-based screening | 15 | 8 | 17 | 8 | 17 |
| No screened | 0 | 50 | 41 | 50 | 41 |
| **2021** |  |  |  |  |  |
| Colonoscopy | 85 | 58 | 58 | 42 | 42 |
| Stool-based screening | 15 | 11 | 22 | 8 | 27 |
| Not screened | 0 | 31 | 20 | 50 | 31 |
| **2022** |  |  |  |  |  |
| Colonoscopy | 85 | 64 | 64 | 64 | 64 |
| Stool-based screening | 15 | 11 | 21 | 11 | 21 |
| Not screened | 0 | 25 | 15 | 25 | 15 |
| **2020 - 2022 average** |  |  |  |  |  |
| Colonoscopy | 85 | 55 | 55 | 49 | 49 |
| Stool-based screening | 15 | 10 | 20 | 9 | 22 |
| Not screened | 0 | 35 | 25 | 42 | 29 |

**Table S3.**
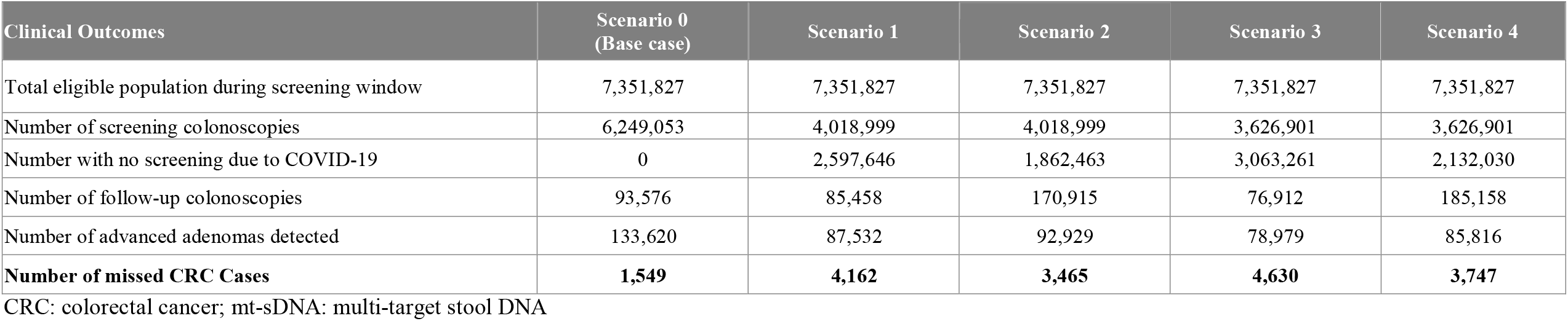
Total 3-year outcomes of screening 45 – 75 year olds for COVID-19 screening scenarios: mt-sDNA.

**Table S4.** Total 3-year outcomes of screening 45 – 75 year olds for COVID-19 screening scenarios: FIT.

| Clinical Outcomes | Scenario 0<br>(Base case) | Scenario 1 | Scenario 2 | Scenario 3 | Scenario 4 |
| --- | --- | --- | --- | --- | --- |
| Total eligible population during screening window | 7,351,827 | 7,351,827 | 7,351,827 | 7,351,827 | 7,351,827 |
| Number of screening colonoscopies | 6,249,053 | 4,018,999 | 4,018,999 | 3,626,901 | 3,626,901 |
| Number with no screening due to COVID-19 | 0 | 2,597,646 | 1,862,463 | 3,063,261 | 2,132,030 |
| Number of follow-up colonoscopies | 40,615 | 41,656 | 83,312 | 37,490 | 90,255 |
| Number of advanced adenomas detected | 130,555 | 85,053 | 87,971 | 76,748 | 80,444 |
| **Number of missed CRC Cases** | **1,662** | **4,237** | **3,615** | **4,698** | **3,910** |

CRC: colorectal cancer; FIT: fecal immunochemical test

**Figure S1.**
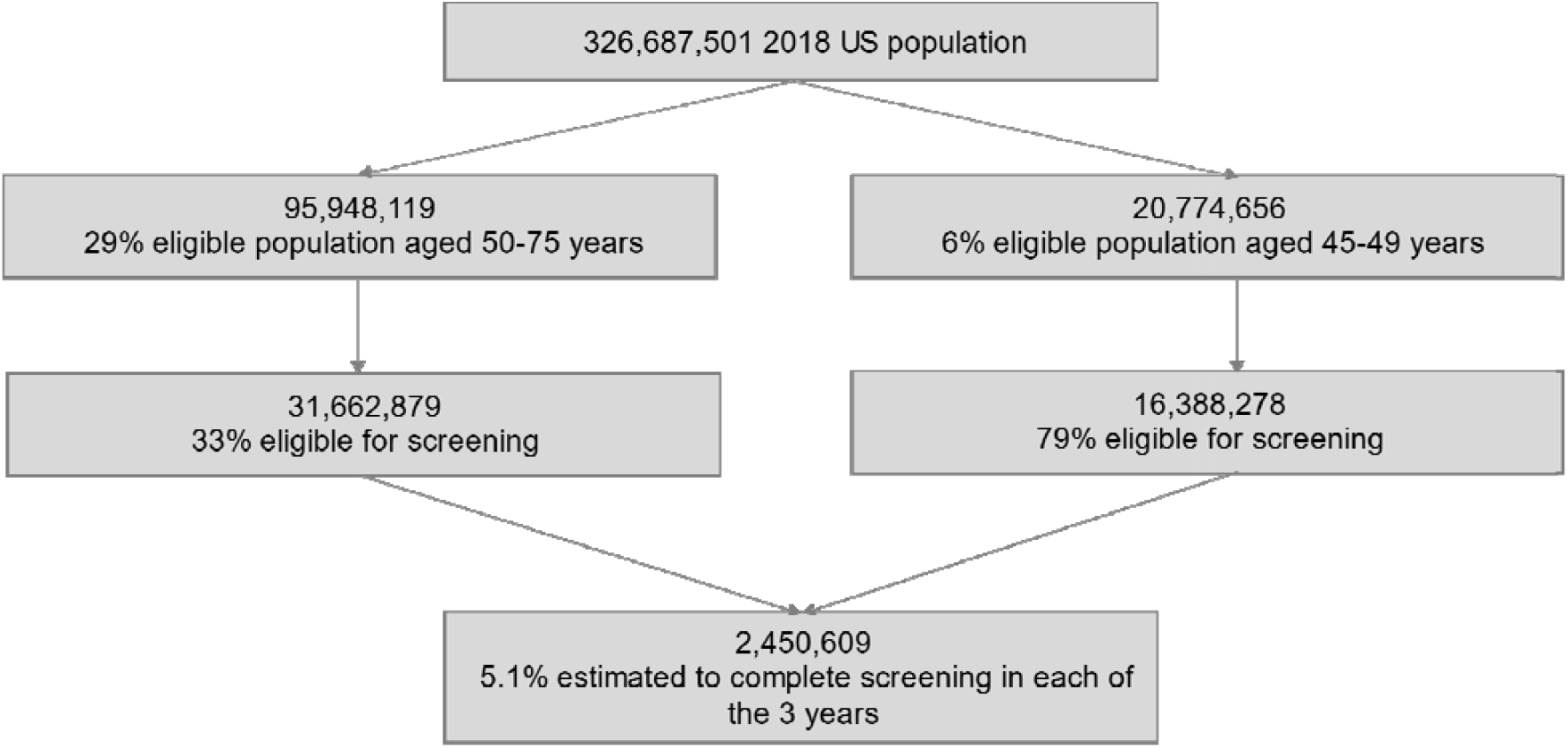
Model patient population flow, sensitivity analysis for people 45 – 75 years of age. US: United States

